# Opportunities to catalyze improved healthcare access in pluralistic systems: a cross-sectional study in Haiti

**DOI:** 10.1101/2020.12.03.20243394

**Authors:** Molly B Klarman, Justin Schon, Youseline Cajusma, Stace Maples, Valery M Beau de Rochars, Chantale Baril, Eric J Nelson

**Affiliations:** Departments of Pediatrics and Environmental and Global Health, University of Florida, Gainesville, FL, USA; Department of Anthropology, University of Florida, Gainesville, Fl; Department of Politics, University of Virginia, Charlottesville, VA; Stanford Geospatial Center, Stanford University, Stanford, CA; Department of Health Services Research, Management and Policy, University of Florida, Gainesville, FL; State University of Haiti, Port-au-Prince, Haiti

## Abstract

**Introduction:** Gains to ensure global healthcare access are at risk of stalling because some old resilient challenges require new solutions. Our objective was to study a pluralistic healthcare system that is reliant on both conventional and non-conventional providers to discover opportunities to catalyze renewed progress.

**Methods:** A cross-sectional study was conducted among households with children less than 5 years of age in Haiti. Households were randomly sampled geographically with stratifications for population density. Household questionnaires with standardized cases (intentions) were compared to self-recall of health events (behaviors). The connectedness of households and their providers was determined by network analysis.

**Results:** A total of 568 households (incorporating 2900 members) and 65 providers were enrolled. Households reported 636 health events in the prior month. Households sought care for 35% (n=220) and treated with home remedies for 44% (n=277). The odds of seeking care increased 217% for severe events (aOR=3.17; 95%CI 1.99-5.05; p< 0.001). The odds of seeking care from a conventional provider increased by 37% with increasing distance (aOR=1.37; 95%CI 1.06-1.79; p=0.016). Despite stating an intention to seek care from conventional providers, there was a lack of congruence in practice that favored non-conventional providers (McNemar’s Chi-squared Test p<0.001). Care was sought from primary providers for 68% (n=150) of cases within a three-tiered network; 25% (n=38/150) were non-conventional.

**Conclusion:** Addressing geographic barriers, possibly with technology solutions, should be prioritized to meet healthcare seeking intentions while developing approaches to connect non-conventional providers into healthcare networks when geographic barriers cannot be overcome.

**Article Summary:**

- The study is inclusive of both conventional and non-conventional healthcare providers, reflecting Haiti’s pluralistic healthcare system.
- The study utilized randomized geospatial sampling method to ensure participants were geographically representative of all families in the study area.
- Employed an unique application of network analysis to observe relationships between families and healthcare providers to identify approaches to increase healthcare access in seemingly change resilient pluralistic systems.
- A limitation of the approach to compare healthcare seeking intentions and behaviors was that the standardized case scenarios did not cover all health event scenarios.
- A limitation on enrollment was that non-conventional healthcare providers were more difficult to locate than conventional providers.

## INTRODUCTION

Improving access to healthcare is one of the highest global health priorities set by the Sustainable Development Goals (SDG) (2015)^1^. SDG 3.8 seeks to “ achieve universal health coverage (UHC)”, however the current rate of progress is insufficient to reach this target by 2030. Low and middle income countries (LMIC) are the furthest off track^2^ ^3^, and UHC tracking indicators show no significant gains for children between 2010 and 2020^4^. The COVID-19 pandemic will exacerbate the limited progress^5^. Innovative approaches are needed to overcome resilient barriers to catalyze progress to achieve universal healthcare access.

Common barriers are accessibility, availability, and acceptability^6^. Determinants inside and outside of households influence when a potential barrier manifests as an actual barrier. One method to prioritize determinants for action is to investigate healthcare seeking behaviors starting with illness recognition (‘is the family member sick?’) and response (‘what should the family do?’). Determinants associated with ‘recognition’ and ‘response’ include biomedical understanding of illness^7^, ability to recognize danger signs^8^, caregiver’s perception of illness severity^9^-^11^, age^12^, level of education^7^ ^8^ and marriage status^11^, number of symptoms^11^, gender^13^ ^14^, rural/urban location^7^ ^15^, intra-household relationships^16^ ^17^, distance^15^ ^18^ ^19^, finances ^17^ ^19^ ^20^ and wait times^18^.

Once a decision is made to seek care, factors that influence provider selection reveal additional barriers. LMIC healthcare systems are poorly defined with a range of conventional and non-conventional providers; alternative terminologies are qualified/unqualified or formal/informal^21^. Parallel and differential access to conventional and non-conventional providers manifests in chaotic systems^22^. Conventional providers can be defined as licensed doctors and nurse practitioners at government or non-governmental organization (NGO) facilities. Non-conventional providers can be defined as traditional healers, medication vendors, unlicensed practitioners and pharmacists. Provider selection exposes conflicts between a patient’s intention (‘would do’) and behavior (‘did do’)^23^ ^24^. These conflicts represent an opportunity to reveal unanticipated solutions to improve access to care.

The associated networks within pluralistic healthcare systems rely on the relationships among and between conventional and non-conventional providers^25^ ^26^. Conventional primary care providers, including community health workers, are assumed to be the first access point into healthcare systems^27^, however non-conventional providers have an important and underappreciated role^28^. Non-conventional providers often practice in parallel without disclosure to conventional providers which creates alternative pathways for seeking care^29^. These complex relationships are not adequately understood, yet are essential to improving healthcare access^30^-^32^.

Haiti was chosen as a generalizable setting to address these knowledge gaps. Access to healthcare in Haiti is low at 23% nationwide and 5% among the rural population^33^. Non-conventional healthcare services and traditional medicine are common^32^ ^34^-^36^. Child health and wellness is substandard with Haiti ranking 150 out of 180 countries^37^. The under five year mortality rate is 67/1,000 live births compared to 41/1,000 live births globally^38^. Healthcare seeking behavior in Haiti has been evaluated for mental health^39^, prenatal care^40^, childbirth location^41^ and cost^42^ ^43^. Our objective was to address actionable knowledge gaps, having generalizable impact beyond Haiti, on care seeking behavior and provider selection from conventional and non-conventional providers, and their associated networks.

## METHODS

### Study design and participants

In this cross-sectional study, consenting participants were enrolled at the household and healthcare provider levels. Household inclusion criteria were households with an adult (18 years or older) head-of-household (HoH) and at least one child under 5 years. Provider inclusion criteria were adult healthcare providers identified by an enrolled household and located within the study area. The 477 sq km study area, encompassing the communes of Gressier and Leogane Haiti, was divided into two-square km grid-cells. Population density was determined by approximating the number of structures within the grid cell using satellite data from the Oak Ridge National Laboratory (USA). Density per grid cell was categorized by structure quartiles; low (0-112 structures), medium (113-397 structures) and high (398-4664 structures). Each grid cell was divided into Thiessen polygons using ArcGIS Online (Esri version 18.0.3) with divisions that reflected the number of households targeted for enrollment; low =18, medium =24, and high =53 per grid cell. The first house encountered in a polygon was recruited. Grid-cells were surveyed sequentially (e.g. low, medium, high) and randomly ordered within each category across a 12-month study period (Aug 2018-July 2019). Although ten grid-cells of each density were designated for survey, six of each were sampled because of logistical limitations. Adult participants provided informed written consent. Ethical approvals were obtained from the Comité National de Bioéthique (National Bioethics Committee of Haiti; 1718-35) and the University of Florida (IRB201703246).

### Data Collection

Data were collected by one enumerator and one nurse. Survey instruments included two thirty-minute in-person questionnaires administered with REDCap mobile version 9.1.1 and were piloted with non-participant households. A household questionnaire collected demographic and socio-economic data, and healthcare seeking behavior for standardized cases and health events (appendix). A health event was defined as any illness a household member experienced in the previous month, regardless of whether care was sought. The standardized respiratory and diarrhoeal cases consisted of hypothetical scenarios involving an ill child at 10 PM with typical symptoms of acute respiratory infection or diarrhoea. A provider questionnaire captured details about the facility/business, personnel qualifications and resources available (appendix). Conventional providers were licensed persons who worked at a licensed facility. For large facilities with greater than 200 patients per month, the facility itself was defined as a ‘provider’. Non-conventional providers were licensed or non-licensed providers at non-licensed facilities, or mobile non-licensed providers.

### Statistical Analysis

Household demographics and health event characteristics were described by proportions for categorical variables and medians with interquartile ranges (IQR) for continuous variables. Social economic status (S.E.S.) index was generated using a summative score of the following components: land, animal, bank account and phone ownership, HoH education level, household electricity source, fuel source, water source, primary source of income, primary transportation method, sanitation type, household floor, wall and roof type, and grid cell density. Provider characteristics were described similarly. The two primary analyses compared (i) households that did and did not seek care for health events and (ii) households that did and did not seek care from a conventional provider. For care-seeking, we used bivariate and multivariate logistic regression models with seeking care as the dependent variable and expressed the results as odds ratios (ORs) and adjusted odds ratios (aORs) for care sought. Similar methods were used to compare the selection of seeking care from a conventional versus non-conventional provider. Health events were analyzed as discrete variables. Two-mode whole networks ^44^ ^45^ of households and providers compared provider types identified/sought in standardized cases and health events; sub-analyses were performed for respiratory infection (‘cough’ or ‘cold’ with fever), diarrhoeal illness (‘diarrhoea’ with/without blood), and “ other” cases (all other illness types not categorized as respiratory or diarrhoea). Ego networks^44^ ^46^ were generated for commonly identified providers in the whole-network analysis. One-mode provider referral networks^44^ were generated. Statistical significance was defined at α=0.05 and 95% confidence intervals are provided. Missingness that was not excluded was for provider types identified by households but not located and enrolled; for regression models, case-wise deletion was used. Analyses were completed in Stata (v11) and the igraph, and in R (R Foundation for Statistical Computing; packages included ‘sf’ by E. Pebesma and R. Bivand 2018)^47^ ^48^.

### Patient and public involvement

Patients or the public were not involved in the design, or conduct, or reporting, or dissemination plans of our research.

## RESULTS

### Household characteristics

A total of 868 households were randomly screened and 568 (65%) were enrolled with a distribution of 112, 144 and 312 between low, medium and high-density grid-cells; 24% were excluded because the household had no children under 5 years (figure 1A, table 1). The average household had 5.2 members, and among the 2900 household members, 58% were under 21 years.

**Table 1.** Household characteristics

| Grid cell density | All<br>N=568 | Low<br>n=112 | Medium<br>n=144 | High<br>n=312 |
| --- | --- | --- | --- | --- |
| Household members: n(%) |  |  |  |  |
| 0 to 1 years | 322 (11) | 63 (11) | 79 (11) | 180 (11) |
| 2 to 4 years | 436 (15) | 76 (14) | 115 (16) | 245 (15) |
| 5 to 10 years | 440 (15) | 78 (14) | 106 (15) | 256 (16) |
| 11 to 20 years | 481 (17) | 89 (16) | 141 (19) | 251 (16) |
| ≥21 years | 1221 (42) | 253 (45) | 284 (39) | 684 (42) |
| Total members | 2900 | 559 (19) | 725 (25) | 1616 (56) |
| Highest-education level (HoH): n(%) |  |  |  |  |
| None | 89 (16) | 26 (23) | 38 (26) | 25 (8) |
| Below secondary | 247 (43) | 42 (38) | 62 (43) | 143 (46) |
| Secondary and above | 232 (41) | 44 (39) | 23 (31) | 144 (46) |
| Transportation: n(%) |  |  |  |  |
| Informal (foot/ bike/ donkey) | 176 (31) | 52 (46) | 64 (44) | 60 (19) |
| Public | 74 (13) | 0 (0) | 1 (1) | 73 (23) |
| Motorcycle taxi | 295 (52) | 55 (49) | 78 (54) | 162 (52) |
| Private car/ motorcycle | 19 (3) | 2 (2) | 1 (1) | 16 (5) |
| Primary source of income: n(%) |  |  |  |  |
| Agriculture/ Animal husbandry | 266 (47) | 92 (82) | 105 (73) | 69 (22) |
| Vendor (Commerce) | 165 (29) | 11 (10) | 24 (17) | 130 (42) |
| Tradesperson | 61 (11) | 5 (4) | 7 (5) | 49 (16) |
| Salaried employment | 35 (6) | 2 (2) | 2 (1) | 31 (10) |
| None | 11 (2) | 0 (0) | 1 (1) | 10 (3) |
| Other | 30 (5) | 2 (2) | 5 (3) | 23 (7) |
| S.E.S index<sup>a</sup>: n(%) |  |  |  |  |
| First quintile | 116 | 59 (53) | 53 (37) | 4 (1) |
| Second quintile | 112 | 29 (26) | 40 (28) | 43 (14) |
| Third quintile | 113 | 20 (18) | 34 (24) | 59 (19) |
| Fourth quintile | 115 | 2 (2) | 12 (8) | 101 (32) |
| Fifth quintile | 112 | 2 (2) | 5 (3) | 105 (34) |

**Figure 1.**
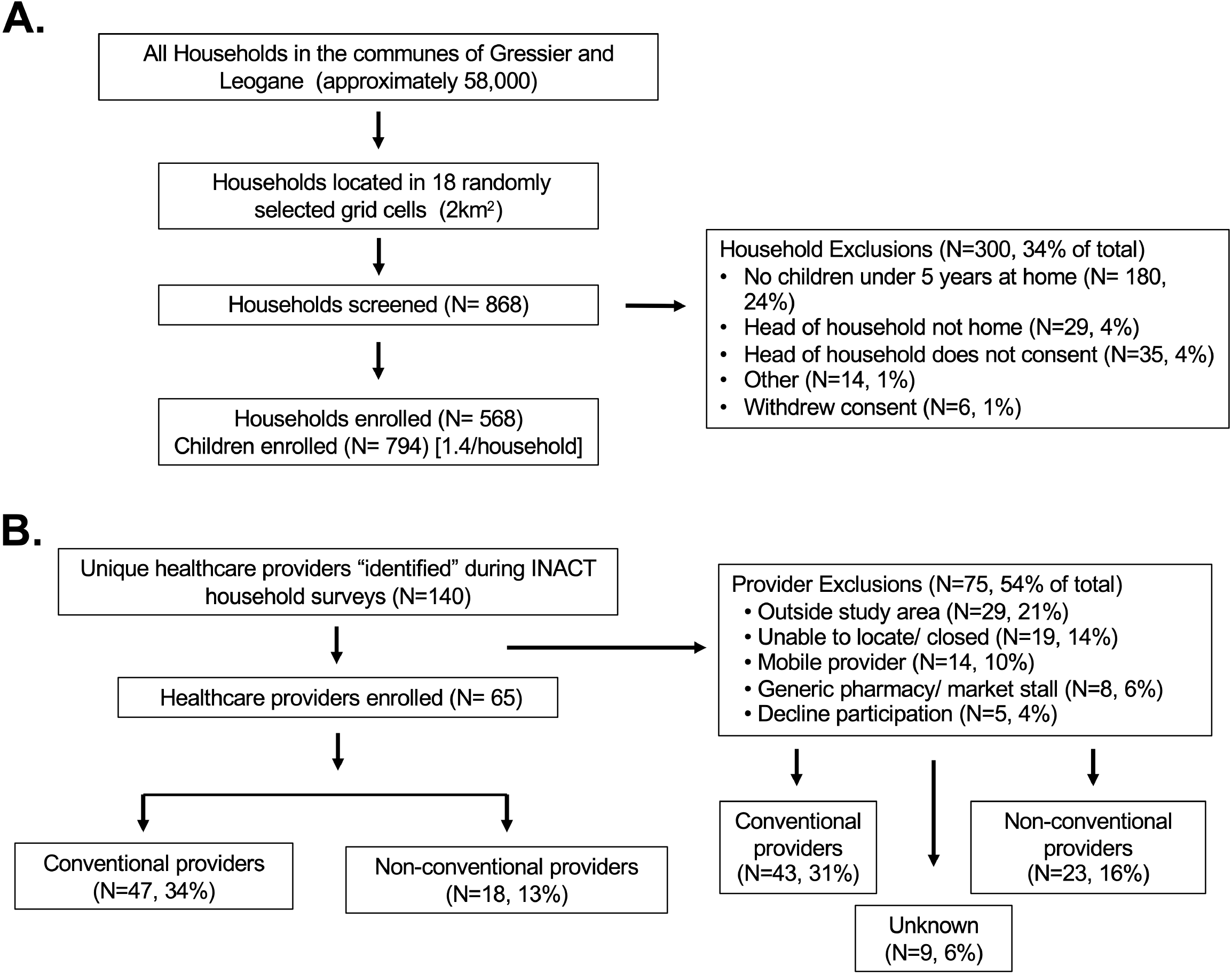
Participant enrollment. **A.** The study enrolled 568 households representing 2900 household members out of 868 households screened. **B.** The study enrolled 65 providers out of 140 identified by households using both standardized case and health event questionnaires; percentages are of those identified by households.

### Health event characteristics

A total of 636 health events were identified (tables 2,S1,S2); 22%, 50%, and 28% of households reported 0, 1, or at least 2 health events in the previous month respectively. The most common symptoms were congestion, fever and cough, present in 61%, 52% and 49% of health events respectively. The most common perceived cause of health events was humoral pathology^49^ (41%), considered an imbalance of ‘hot’ and ‘cold’ within the body caused by environmental exposures.

**Table 2.**
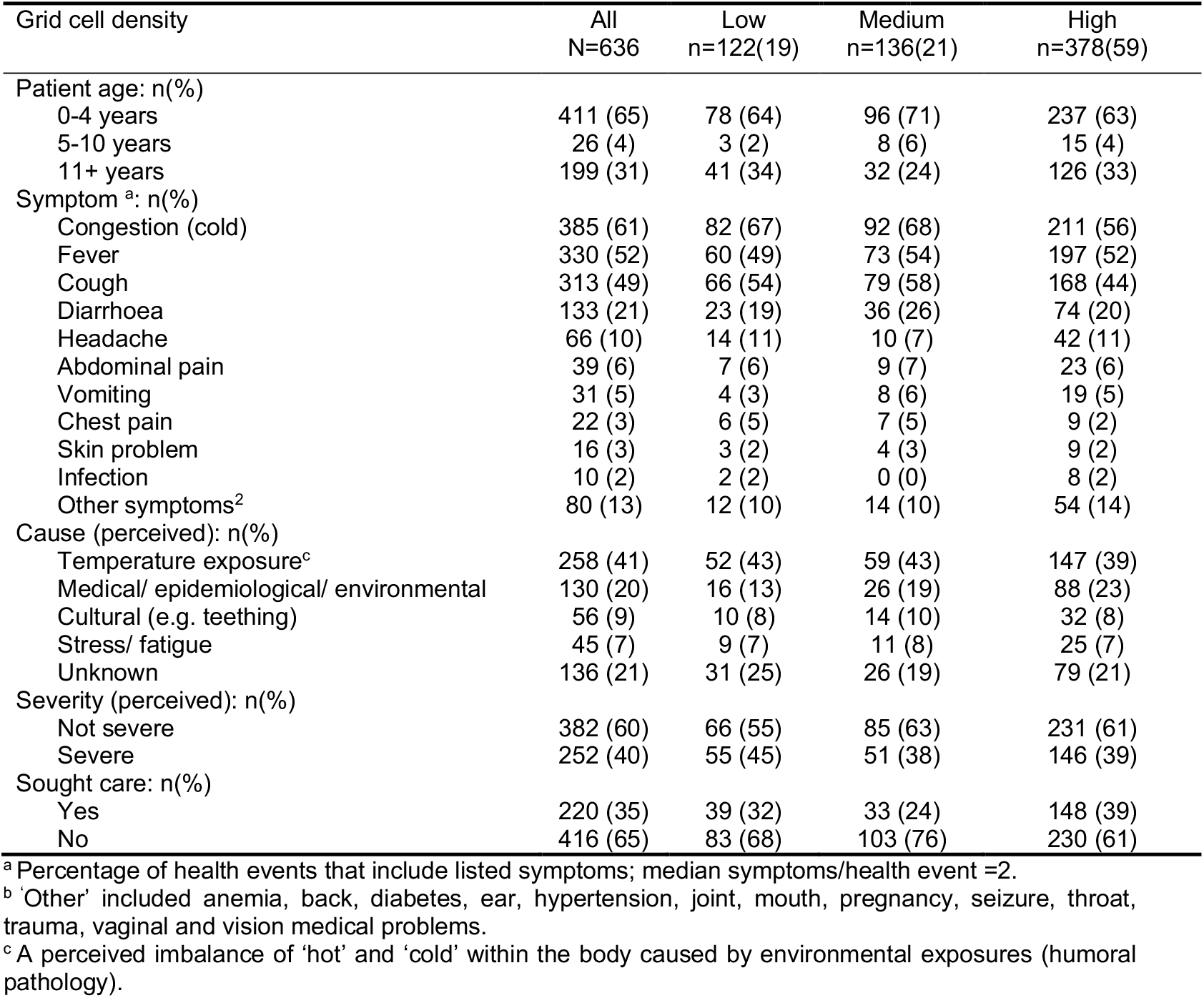
Health event characteristics

**Table 3.** Determinants to seek care for health events

|  | Care sought<br>n=220 | Care not sought<br>n=416 | OR (CI) | p value | aOR (CI) | p value |
| --- | --- | --- | --- | --- | --- | --- |
| Patient age: n(%) |  |  |  |  |  |  |
| 0-4 years | 131 (60) | 280 (67) | 0.71 (0.51-1.00) | 0.052 | Ref | Ref |
| 5-10 years | 10 (5) | 16 (4) | 1.19 (0.53-2.67) | 0.672 | 0.77 (0.22-2.63) | 0.675 |
| 11+ years | 79 (36) | 119 (29) | 1.38 (0.98-1.96) | 0.068 | 0.75 (0.40-1.38) | 0.355 |
| Illness<sup>a</sup>: n(%) |  |  |  |  |  |  |
| Respiratory | 96 (44) | 222 (53) | **0.67 (0.48-0.94)** | **0.018** | 0.75 (0.40-1.42) | 0.378 |
| Diarrhoea | 17 (8) | 35 (8) | 0.91 (0.50-1.66) | 0.757 | 0.69 (0.27-1.73) | 0.426 |
| Respiratory and diarrhoea | 19 (9) | 62 (15) | **0.54 (0.31-0.93)** | **0.025** | 0.99 (0.37-2.65) | 0.980 |
| Other | 88 (40) | 96 (23) | **2.22 (1.56-3.15)** | **<0.001** | Ref | Ref |
| Distance (km)<sup>b</sup>: median (IQR) | 3.2 (1.3-5.5) | 3.5 (1.5-6.1) | 0.97 (0.93-1.02) | 0.239 | 0.96 (0.91-1.01) | 0.135 |
| Severity (perceived): n(%) |  |  |  |  |  |  |
| Not-Severe | 92 (42) | 290 (70) | Ref | Ref | Ref | Ref |
| Severe | 127 (58) | 125 (30) | **3.20 (2.28-4.50)** | **<0.001** | **3.17 (1.99-5.05)** | **<0.001** |
| Time of health event<sup>c</sup> |  |  |  |  |  |  |
| Day | 101 (48) | 138 (36) | Ref | Ref | Ref | Ref |
| Night | 108 (52) | 242 (64) | **0.69 (0.50-0.96)** | **0.029** | 0.80 (0.49-1.29) | 0.359 |
| Transportation: n(%) |  |  |  |  |  |  |
| Informal (foot/ bike/ donkey) | 43 (20) | 130 (31) | **0.53 (0.36-0.79)** | **0.002** | 0.78 (0.28-2.19) | 0.638 |
| Public/ motorcycle taxi | 166 (76) | 267 (64) | **1.72 (1.19-2.49)** | **0.004** | Ref | Ref |
| Private car/ motorcycle | 10 (5) | 17 (4) | 1.12 (0.50-2.48) | 0.785 | 0.49 (0.15-1.58) | 0.235 |
| Highest-education level (HoH): n(%) |  |  |  |  |  |  |
| None | 22 (10) | 66 (16) | **0.59 (0.35-0.98)** | **0.043** | Ref | Ref |
| Below secondary | 88 (40) | 181 (44) | 0.87 (0.62-1.21) | 0.394 | 1.34 (0.60-3.02) | 0.475 |
| Secondary and above | 110 (50) | 169 (41) | **1.46 (1.05-2.03)** | **0.024** | 1.61 (0.68-3.77) | 0.277 |
| Primary source of income n(%) |  |  |  |  |  |  |
| Agriculture/Animal husbandry | 87 (43) | 182 (48) | 0.85 (0.61-1.20) | 0.364 | 5.04 (0.57-44.51) | 0.146 |
| Vendor (Commerce) | 71 (35) | 134 (35) | 1.02 (0.72-1.46) | 0.897 | 2.56 (0.29-22.67) | 0.398 |
| Tradesperson | 33 (16) | 47 (12) | 1.41 (0.87-2.29) | 0.160 | 2.68 (0.29-25.18) | 0.388 |
| Salaried employment | 16 (8) | 25 (6) | 1.24 (0.65-2.38) | 0.512 | 2.95 (0.30-28.67) | 0.351 |
| Grid cell population density: n(%) |  |  |  |  |  |  |
| Low | 84 (32) | 39 (10) | 0.86 (0.57-1.32) | 0.498 | Ref | Ref |
| Medium | 33 (12) | 103 (28) | **0.54 (0.35-0.83)** | **0.005** | 1.11 (0.52-2.35) | 0.783 |
| High | 148 (56) | 230 (62) | **1.66 (1.18-2.34)** | **0.004** | 1.69 (0.82-3.47) | 0.154 |
| Health events per prior month:<br>median (IQR) | 1 (1-2) | 1 (1-2) | 1.08 (0.88-1.32) | 0.458 | 0.94 (0.72-1.23) | 0.666 |
| S.E.S index<sup>d</sup> median (IQR) | 3 (2-4) | 3 (2-4) | **1.45 (1.03-2.03)** | **0.034** | (omitted) | (omitted) |

Forty percent of health events were considered severe. Half of all health events started at nighttime (55%), of these, 36% were severe (table S1). Respiratory infection symptoms started more often at nighttime (66%) compared to diarrhoea that more often started during the daytime (65%) (table S2).

### Determinants to seek care for health events

Healthcare was sought outside the household for 35% (n=220/636) of health events, and most often for children between 0-4 years (65%). The most common reason cited for not seeking care was preference to treat at the household with home remedies (67%; n=277/416) followed by the health event did not necessitate treatment (24%; n=99/416). The majority of households delayed seeking care (67%; n=138/201), most commonly citing monetary and transportation (46%; n=63/138) barriers. Among those who sought care without delay, 68% (n=43/63) chose to do so because of severity. In the bivariate analysis, decreased odds of seeking care was associated with nighttime events (OR=0.69; 95%CI 0.50-0.96; p=0.029) and informal transport (OR=0.53; 95%CI 0.36-0.79; p=0.002). Increased odds of seeking care were associated with illness severity (OR=3.20; 95%CI 2.28-4.50; p< 0.001), high density grid-cells (OR=1.66; 95%CI 1.18-2.34; p=0.004) and high S.E.S. (OR=1.45; 95%CI 1.03-2.03; p=0.034). In the multivariate analysis, the odds of seeking care increased by 217% with severe illness (aOR=3.17; 95%CI 1.99-5.05; p<0.001).

### Provider characteristics

Households identified 140 providers and 65 (46%) were located and enrolled (figure 1B; table S3). Failure to enroll providers was most commonly due to the provider being located outside the study area (21%; n=29) or an inability to locate the provider/ had ceased operation (14%; n=19). Details from the household surveys enabled the determination of provider type and setting (table S3). Among surveyed providers, 68% (n=44) had a business license. Variation was observed by grid cell density; only 4 providers were identified in the low-density grid-cells and none were available after 8 pm (table S3).

### Determinants to seek care from a conventional provider

Care was sought from a conventional provider for 83% (n=180/218) and a non-conventional provider for 17% (n=38/218) of health events (table S4). Bivariate analysis found decreased odds of seeking care from a conventional provider was associated with informal transport (OR=0.33; 95%CI 0.15-0.71; p=0.005) and HoH with no formal education (OR=0.32; 95%CI 0.12-0.82; p=0.018). The odds increased in association with S.E.S. (OR=2.78; 95%CI 1.25-6.25; p=0.12). In multivariate analysis, increased odds of seeking care from a conventional provider was associated with increased travel distance (aOR=1.37; 95%CI 1.06-1.79; p=0.016) and HoH education at or above secondary school (aOR=5.00; 95%CI 1.05-25.00; p=0.044). These findings are consistent with responses that 53% of families (n=116/220) cited distance as the most important factor in provider selection. Households that cited distance traveled a median of 1.5km (IQR 0.7km-3.9km) in comparison to 3.6km (IQR 1.5km-7.8km) for households that did not cite distance. Households traveled further (Mann Whitney, p<.0001) to seek care from conventional providers (median 2.6km, IQR 1.1km-5.9km) than non-conventional providers (median 0.9km, IQR 0.0km-2.3km). The difference in cost was less when households sought care from non-conventional providers (median US $1.28, IQR US $0.16-$2.56) compared to when households sought care from conventional providers (median US $4.49, IQR US $1.92-$7.85; Mann Whitney, p < 0.0001).

### Congruence between care seeking intentions and behaviors by provider type

Comparisons of intentions versus behaviors for the type of provider sought was conducted using data from the household standardized cases (‘where would you go?’) and the household health events (‘where did you go?’). Among all health event types, there was a lack of congruence between intentions and behaviors (OR =0.15; 95%CI 0.04-0.43; p<0.001); 27 of the 31 non-congruent events were attributed to households that intended to seek care from a conventional provider but switched to non-conventional (table S5). In bivariate analysis, ARI with diarrhoea and informal transportation were associated with increased non-congruence. In contrast, HoH education level of secondary and above and high-density grid-cells were associated with congruence. Multivariate analysis found no significant factors (table S6).

### Network analysis of care-seeking intentions versus behaviors

Two-mode whole networks were used to evaluate the relationships between households and providers identified through standardized cases and health events (figure 2). The number of households linked to each provider per health event was statistically different compared to standardized respiratory (Mann Whitney, p=.014) and diarrhoeal (Mann Whitney, p=.007) cases; distance traveled was statistically different compared to diarrhoeal cases (Wilcoxon, p=.001); sub-analysis comparisons for respiratory and diarrhoeal specific health events were similar (figure S1). Two-mode ego network analyses focused on a public clinic (C1) and a private hospital (H1) commonly identified in the whole-network analysis as the intended provider (figure 2); in practice, households sought care from these providers less often (Student *t*-test; both p<0.001). In one-mode analysis, providers identified 33 conventional and 0 non-conventional referral providers that formed a three-tiered network (figure. S2A); 45% (n=15) received one referral at the lowest tier, 42% (n=14) received between 2-10 referrals at the middle tier and 12% (n=4) received 10 or more referrals at the highest tier. Sixty-eight percent of households sought care from providers receiving zero referrals. Referral networks from standardized respiratory and diarrhoeal cases were similar (figure S2B,C).

**Figure 2.**
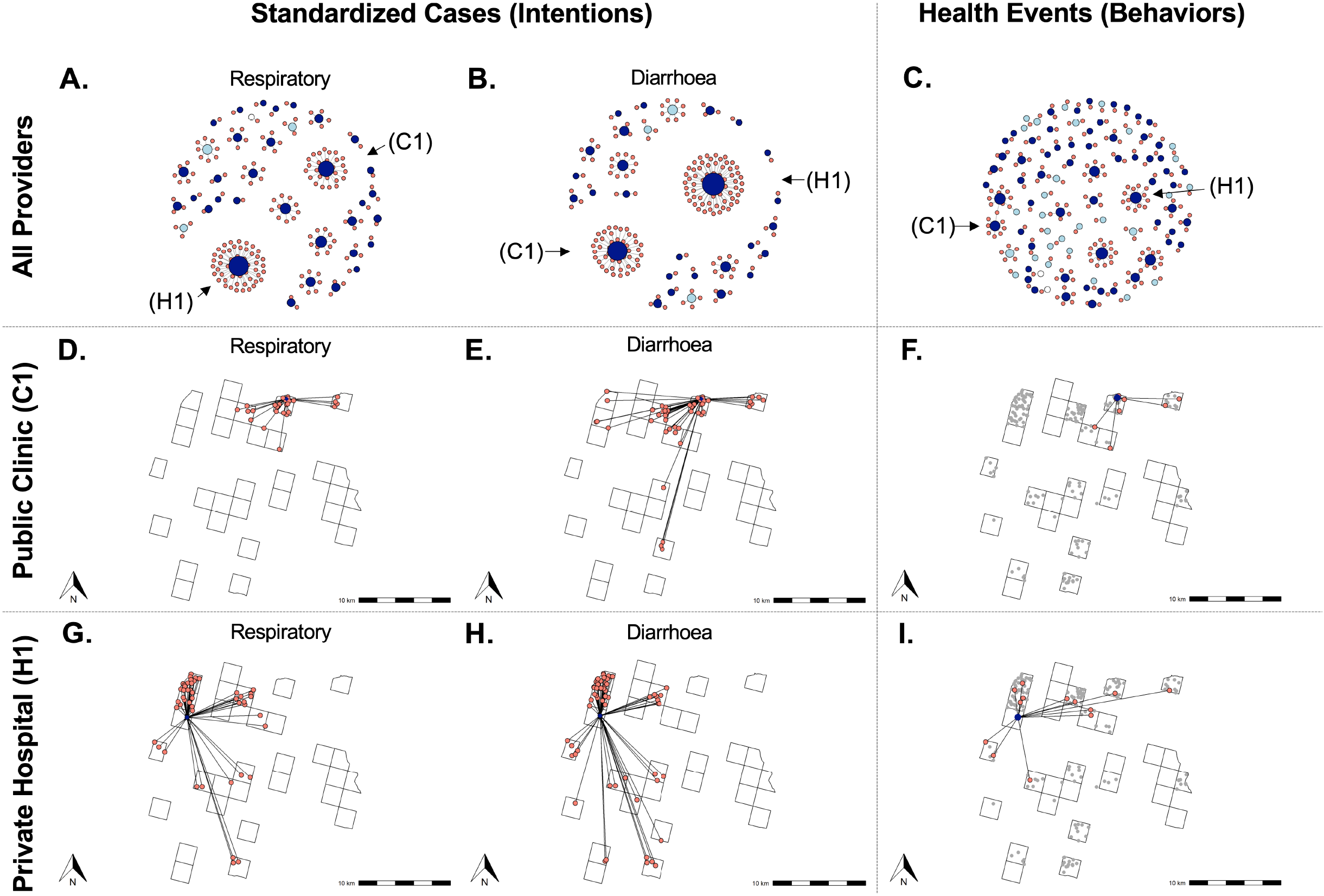
Provider selection. Among households with health events in which care was sought (n=180), two-mode whole network analysis was used to elicit care-seeking intentions for standardized respiratory cases (**A)**, standardized diarrhoeal cases (**B)**, and health events **(C)**. The median number of household-provider linkages per provider and distance traveled were 2 (IQR=1-5) and 3.2km (IQR=1.3km-5.6km), 2.5 (IQR=1-5) and 4.0km (IQR=1.9km-6.4km) and 1 (IQR=1-2) and 2.1km (IQR=1.0km-5.4km) for respiratory standardized cases (A), diarrhoeal standardized cases (B) and health events (C), respectively. Ego network analysis of the two most commonly identified providers. Households that selected public clinic C1 in the respiratory standardized case (n=26; **D**), the diarrhoeal standardized case (n=50; **E**), and/or sought care at C1 for a health event (n=9; **F**). Households that selected private hospital H1 in the respiratory standardized case (n=51; **G**), the diarrhoeal standardized case (n=64; **H**), and/or sought care at H1 for a health event (n=13; **I**).

## DISCUSSION

Should care be sought, and from whom, are fundamental questions when medical problems arise in households. The decisions may trigger entry into a linear healthcare system with primary, secondary and tertiary conventional providers or a pluralistic system that includes non-conventional providers. In this cross-sectional study, we found that approximately one-third of health events were not treated, one-third were treated at the household and one third sought care. When care was sought, it was for severe illnesses and from a mixture of conventional and non-conventional providers, largely disconnected from healthcare networks, despite intentions to seek care from connected conventional providers. The findings reveal insights on how to catalyze improved healthcare access in seemingly change resilient healthcare settings.

Households living in low population density regions had the lowest S.E.S. compared to those in high population density regions. Households approached health events with a pragmatic mindset aware of healthcare access barriers. Barriers to seeking care included nighttime presentation, limited access to motorized transport, and poverty. Disease severity was the major determinant to seek care. Families delayed seeking care because of financial and transport barriers, unless the event was severe or not resolving. These data document that poverty and geographic isolation in rural communities are barriers to seek care.

Providers were identified by households in both the standardized cases and the health events. Increased S.E.S correlated with seeking care from a conventional provider. Multivariate analysis identified increased distance, education, and higher population density as correlates of seeking care from a conventional provider. These findings highlight that once a household decides to seek care, poverty and geographic isolation again influence the decision to seek care from a conventional versus non-conventional provider.

Intentions (‘would do’) and behaviors (‘did do’) for care-seeking and provider selection were compared by analyzing standardized cases against the health events. Conflict between household intentions and behaviors for seeking care from a conventional provider was found. The directionality was from an intention to seek care from a conventional provider to care sought from a non-conventional provider. These analyses highlight the inequity that both poor and geographically isolated households confront between intentions and behaviors. The data also suggest that poor and isolated households are not content with the choice they are forced to make. Network analysis compared intentions versus behaviors for provider selection. A stark contrast was revealed between an intention to seek care from commonly shared providers yet a behavior to seek care from providers that were less commonly shared. For example, networks analysis of two highly desired conventional providers (C1; H1) found that in practice, less care was sought from these providers.

Disparity between intentions and behaviors can be found broadly across the domain of healthcare. However, this study provides insight that when faced with poverty and isolation, households are forced to turn to nearby providers that are often non-conventional. Methods to reduce non-congruence between intentions and behaviors are needed. We advocate that governmental and non-governmental organizations develop initiatives to investigate how to link disconnected non-conventional providers into the healthcare networks with conventional providers. In parallel, households may desire and need an innovative alternative mechanism to ‘bypass’ barriers they face when attempting to seek care from a conventional provider. A new model (figure 3) may include improved access to centralized services, or extending access to/near households by mobile healthcare services either physically or electronically through telemedicine.

**Figure 3.**
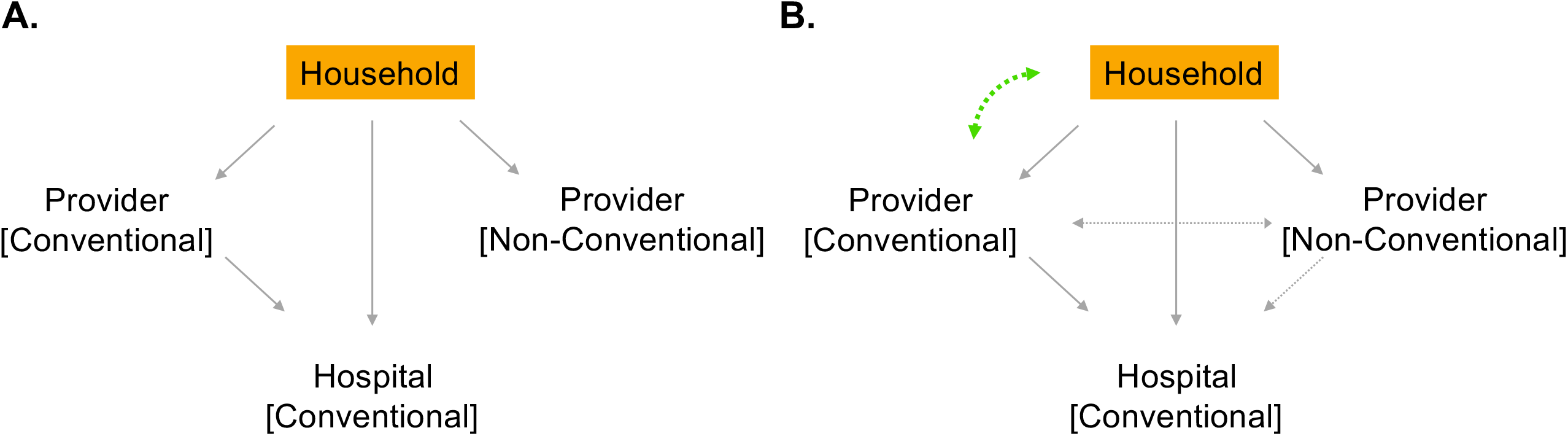
Model to improve access to healthcare in a pluralistic system. A. Current relationships between households and providers. B. Opportunities to improve access to care by better connecting households to conventional providers, and non-conventional providers to the healthcare network. Solid gray arrows represent current connections. Dashed green curved arrow represents a bypass mechanism to improve access to conventional primary providers by innovative mechanisms (e.g. dedicated transport/assistance, mobile outreach clinics, telemedicine). Dashed gray arrows represent connections to link conventional/non-conventional providers.

These findings should be viewed within the context of the study limitations. First, the standardized case questionnaires were designed *a priori* to investigate the decision-making process to seek care for children with respiratory or diarrhoeal disease at night. The analytic strategy to compare standardized cases as intentions and health events as behaviors was established *post-hoc*. The limitation is that the specific standardized cases were limited to children who developed symptoms at night which reduced the strength of comparison to the diversity of health events despite sub-analysis to compare cases by chief complaint. The sample size of health events with only respiratory or diarrhoeal complaints alone also limited the strength of comparisons with standardized cases. Second, adjustments for household clustering were not performed because 72% of households had zero or one health event. Third, providers identified by households were difficult to locate and only a portion of providers were located. This resulted in missingness and an information bias such that mobile drug vendors were interviewed less. Lastly, there was a risk of recall bias because questionnaires relied on one-month and one-week recall by households and providers, respectively. Despite these limitations, the approach and findings represent a meaningful contribution.

### Conclusion

The study supports a model that households are likely to treat illnesses at home without assistance. When care is sought, it is for severe illnesses and often from a mixture of disconnected conventional and non-conventional providers despite an intention to seek care from a conventional provider. The findings support bridging geographic barriers, possibly with technology solutions, to meet healthcare seeking intentions while developing approaches to connect non-conventional providers into healthcare networks when geographic barriers cannot be overcome.

## Data Availability

Data is available at the Dryad data repository.

## Contributions

EJN and MBK conducted the literature search. The study was designed by EJN and MBK. MBK and CY implemented the study and collected data with guidance from SM. SM provided the approach and tools for geospatial mapping. Data analysis, interpretation, figure design, and writing was conducted by JS, MBK and EJN. Clinical oversight in Haiti was governed by CB with assistance by MVBDR. Communication with IRBs was performed by CB, MVBDR and EJN. IRB documentation was written and managed by MBK and EJN.

## Acknowledgements

We thank the households and providers that agreed to participate in this study, and diligent field research team who made this study possible. We are grateful to R. Autrey, K. Berquist, D. Schatz and S. Rivkees at the University of Florida for their support and helpful discussions. We thank M. Gurka, R. Vacca and C. McCarty at the University of Florida for guidance and insight on study design and social network analysis. We also express our gratitude to the Christianville Foundation (Gressier, Haiti) for their shared commitment to healthcare access and providing infrastructure for implementation.

## Data availability

Data are available in the Supplementary Appendix.

## Disclaimer

These funders had no role in study design, data collection and analysis, decision to publish, or preparation of the manuscript.

## SUMMARY OF ADDED BENEFIT AND CLINICAL IMPACT

To best describe what is known about the subject in this manuscript, we performed two searches in PubMed for reports published after January 1, 2010 in all languages. The first search consisted of the search terms [Haiti] AND [care seeking] OR [health seeking] OR [healthcare seeking]. The criteria identified 16 publications. Four of the articles had a primary focus of healthcare seeking behaviors in Haiti; 1 pertained to rabies, 2 pertained to mental health and 1 pertained to equitable access to healthcare. Care seeking in Haiti was a component in an additional 6 articles, although not as the primary research objective; 1 pertained to rabies, 1 pertained to cholera patients, 1 pertained to suicide, 1 multi-country study pertained to child health and family planning, 1 multi-country study pertained to user fees and 1 multi-country study pertained to malaria and the private health sector. The remaining 6 articles did not pertain to healthcare seeking in Haiti.

The second search consisted of the terms “ network analysis” AND “ healthcare providers” OR “ healthcare provider” OR “ referral”. The criteria identified 81 publications. The results were filtered to remove articles with the terms “ United States”, U.S., Europe, Australia, “ neural network analysis”, and “ thematic network analysis” and non-primary research results were also removed. This yielded 20 publications. Three papers used network analysis to investigate the roll of peer networks, one pertaining to diet and exercise in Korea, one pertaining to risk of migraines in Taiwan, and one pertaining to theoretical support programming for cancer patients. Two papers used network analysis to characterize professional advice seeking behavior; one among physicians in Pakistan and the other among primary healthcare workers in Ethiopia. Three papers used network analysis to assess health systems functioning, one pertaining to communication during hospital patient discharge in the US, one pertaining to patient flow in a US hospital and one pertaining to health systems integration in China. One paper created referral networks among community healthcare providers and their patients 65 and older in the United Kingdom to better inform the organization of community services. Another paper from the United Kingdom used network analysis to map and understand naturally occurring communities that could assist in the development of a new primary care network initiative. Three papers examined relationships among networks of organizations who support the health of specific populations, one pertaining to organizations involved in cancer screening of South Asians in Canada, one pertaining to professionals who provide psychosocial care to victims of violence in Colombia, and one pertaining to organizations who address unmet social needs and their clinical partners in the US.

Despite this body of literature, we did not identify studies that used original data to investigate general healthcare seeking behavior in Haiti. Beyond Haiti, we did not identify studies that performed a network analysis to investigate relationships between households and the healthcare providers they frequent, especially in a global health context. Referral networks were represented in the literature but in the context of high-income countries. Therefore, the knowledge this manuscript contributes on pluralistic healthcare systems are of great value globally. The innovative methods of leveraging standardized cases to gauge intentions and the network analysis framework benefit teams faced with similar challenges. The study revealed unanticipated opportunities that should be further evaluated outside of Haiti and considered by international stakeholders as avenues to catalyze renewed SDG progress.

