## Supplementary Appendix for "Opportunities to catalyze improved healthcare access in pluralistic systems: a cross-sectional study in Haiti"

**Dataset (Household and Provider) included separately**

**Figure S1.** Two-mode whole network analysis of care-seeking intentions versus behavior for among households with respiratory and diarrhoeal cases health events.

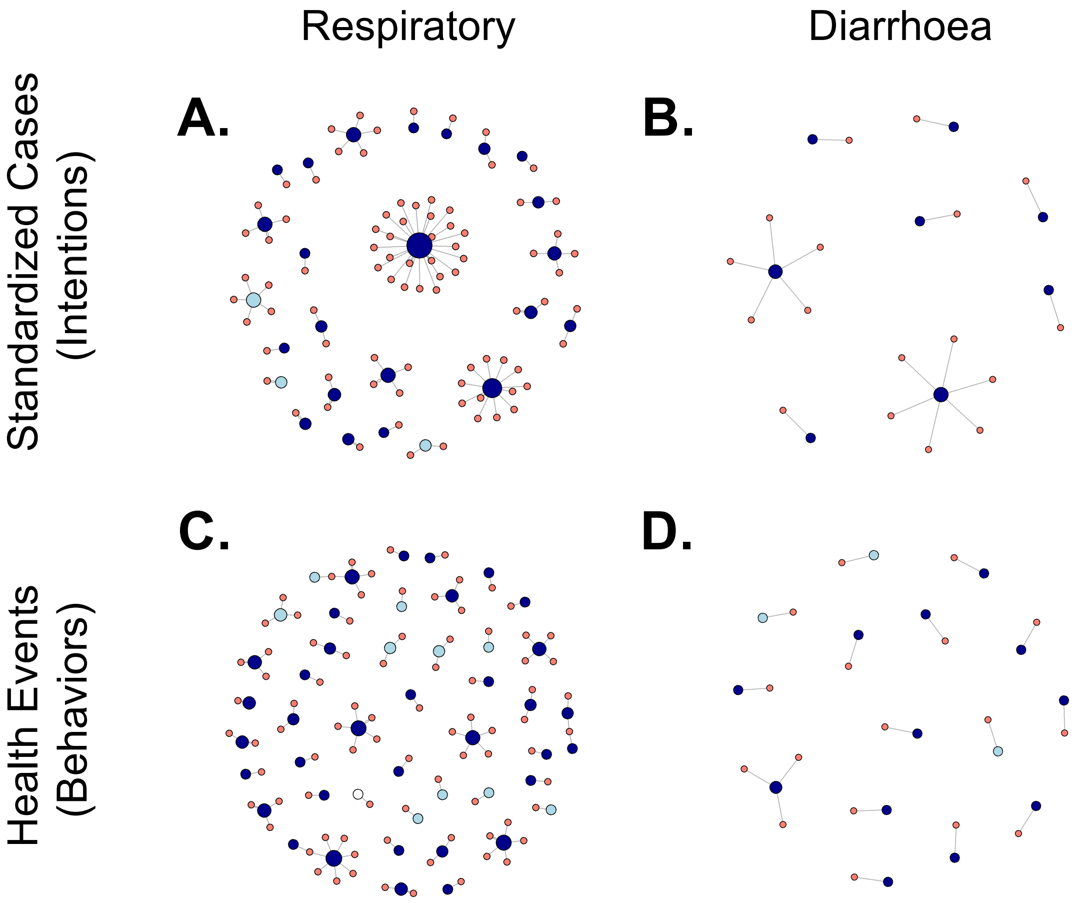

**Figure S1.** Two-mode whole network analysis of care-seeking intentions versus behavior for households that sought care for respiratory or diarrhoeal cases health events. **A.** Standardized respiratory cases among households with respiratory health events (n=96). **B.** Standardized diarrhoeal cases among households with diarrhoeal health events (n=17). **C.** Households with respiratory heath events (n=96). **D.** Households with diarrhoeal heath events (n=17). Salmon circles = households. Dark blue circles = conventional provider. Light blue circles = non-conventional provider. Enumerations are provided (see Appendix table of contents).

**Figure S2.** One-mode provider referral network generated from provider surveys.

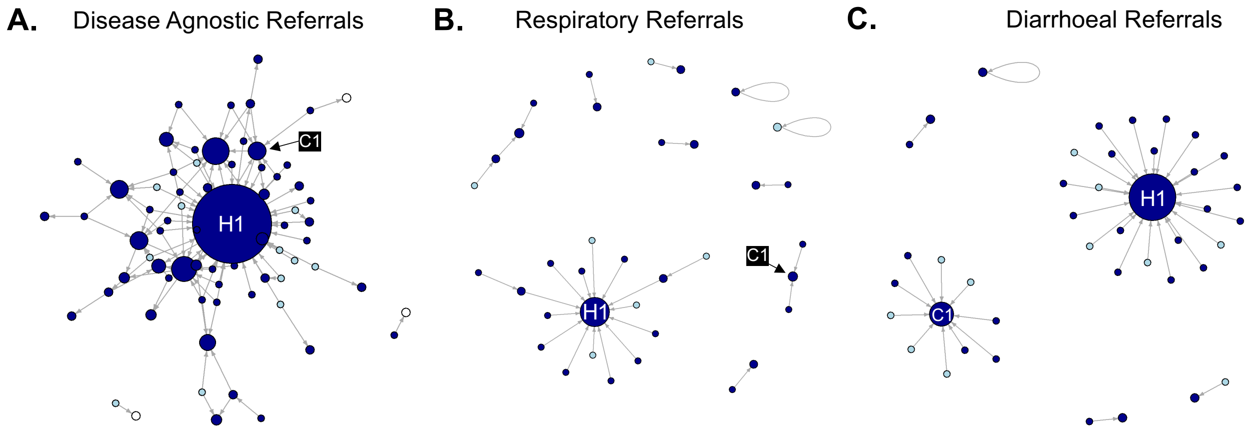

**Figure S2.** One-mode provider referral network generated from provider surveys. **A.** Referral providers identified in the survey question to name up to three referral providers. **B.** Referral providers identified in the respiratory standardized case. **C.** Referral providers identified in the diarrhoeal standardized case. Dark blue and light blue circles represent conventional and non-conventional providers, respectively. Providers of unknown type are represented by clear circles. Gray arrows represent the direction of the referral.

**Table S1.** Distribution of health events by severity**.**

| **Table S1. Distribution of health events by severity** | Total N=636 | Severe health event n=252 (40%) | Not severe health event n=382 (60%) |
|---|---|---|---|
| Time of health event[a]: n(%) | | | |
| Day | 286 (45) | 126 (44) | 158 (55) |
| Night | 350 (55) | 126 (36) | 224 (64) |
| Perceived health event etiology | | | |
| Temperature exposure[b] | 258 (41) | 77 (30) | 180 (70) |
| Medical/epidemiological/environment | 130 (20) | 70 (54) | 59 (46) |
| Cultural (e.g. teething) | 56 (9) | 9 (16) | 47 (84) |
| Stress/fatigue | 45 (7) | 20 (44) | 25 (56) |
| Unknown | 136 (21) | 71 (52) | 65 (48) |
| Provider sought[a]: n(%) | | | |
| Daytime care sought | 204 (96) | 120 (59) | 83 (41) |
| Nighttime care sought | 8 (4) | 5 (63) | 3 (38) |
| Total care sought | 220 (35) | 127 (58) | 92 (42) |
| Total care not sought | 416 (65) | 125 (30) | 290 (70) |
| Grid cell density: n(%) | | | |
| Low | 122 (19) | 55 (45) | 66 (54) |
| Medium | 136 (21) | 51 (38) | 85 (63) |
| High | 378 (59) | 146 (39) | 231 (61) |

[a] Day is between 6AM- 6PM and night is between 6PM-6AM
[b] A perceived imbalance of ‘hot’ and ‘cold’ within the body caused by environmental exposures (humoral pathology)

**Table S2.** Distribution of respiratory and diarrhoeal health events by time and population density

**Table S2. Distribution of respiratory and diarrhoeal health events by time and population density**

| | All events N=636 | Respiratory n=318 | Diarrhoea n=52 | Respiratory and Diarrhoea n=81 |
|---|---|---|---|---|
| Time of health event[a]: n(%) | | | | |
| Day | 286 (45) | 107 (34) | 34 (65) | 29 (36) |
| Night | 350 (55) | 211 (66) | 18 (35) | 52 (64) |
| Grid cell density: n(%) | | | | |
| Low | 122 (19) | 69 (57) | 8 (7) | 15 (12) |
| Medium | 136 (21) | 65 (48) | 8 (6) | 28 (21) |
| High | 378 (59) | 184 (49) | 36 (10) | 38 (10) |

[a] 'Day' is between 6AM- 6PM and night is between 6PM-6AM.

**Table S3.** Provider characteristics

**Table S3. Provider characteristics**

| | All[a] N=140 | Surveyed[b] N=65 | Low[b] n=4 | Medium[b] n=15 | High[b] n=46 |
|---|---|---|---|---|---|
| Type of practice: N(%) | | | | | |
| Conventional | 89 (64) | 47 (72) | 3 (75) | 7 (47) | 37 (80) |
| Pharmacist | 6 (7) | 3 (6) | 0 (0) | 1 (14) | 2 (5) |
| Nurse | 7 (8) | 6 (13) | 1 (33) | 1 (14) | 4 (11) |
| Doctor | 5 (6) | 2 (4) | 0 (0) | 0 (0) | 2 (5) |
| Mix (doctor, nurse, pharmacist) | 71 (80) | 36 (77) | 2 (67) | 5 (71) | 29 (78) |
| Non-conventional | 41 (29) | 18 (28) | 1 (25) | 8 (53) | 9 (20) |
| Other/ Unknown | 10 (7) | 0 (0) | 0 (0) | 0 (0) | 0 (0) |
| Setting: n(%) | | | | | |
| Transient vendor | 18 (13) | 1 (2) | 0 (0) | 0 (0) | 1 (2) |
| Home | 17 (12) | 10 (15) | 0 (0) | 6 (40) | 4 (9) |
| Pharmacy | 13 (9) | 7 (11) | 0 (0) | 1 (7) | 6 (13) |
| Office | 23 (16) | 15 (23) | 3 (75) | 4 (27) | 8 (17) |
| Clinic | 34 (24) | 26 (40) | 1 (25) | 4 (27) | 21 (46) |
| Hospital | 22 (16) | 4 (6) | 0 (0) | 0 (0) | 4 (9) |
| Other/ Unknown | 13 (9) | 2 (3) | 0 (0) | 0 (0) | 2 (4) |
| Features: n(%) | | | | | |
| Business license | | 44 (68) | 3 (75) | 8 (53) | 33 (72) |
| Privately operated | | 51 (78) | 3 (75) | 14 (93) | 33 (72) |
| Signage | | 44 (68) | 3 (75) | 5 (33) | 36 (78) |
| Hours of operation: n(%) | | | | | |
| Open at night[c] | | 14 (22) | 0 (0) | 2 (13) | 12 (26) |
| Open on weekends | | 26 (40) | 1 (25) | 2 (13) | 20 (43) |
| Patient visits/ provider/ week: Median (IQR) | | 38 (14-75) | 32 (29-34) | 27 (5-47) | 43 (15-100) |

[a] Includes all providers identified by households (inside and outside of study area).
[b] Includes only providers surveyed.
[c] Night is defined as 8PM.

**Table S4.** Determinants of selecting a conventional provider for a health event

**Table S4. Determinants of selecting a conventional provider for a health event**

| | Conventional n=180* | Non-conventional n=38* | OR (CI)* | P* | aOR (CI)* | P* |
|---|---|---|---|---|---|---|
| Patient age: n(%) | | | | | | |
| 0-4 years | 110 (63) | 20 (53) | 1.41 (0.70-2.86) | 0.334 | Ref | Ref |
| 5-10 years | 9 (5) | 1 (3) | 1.96 (0.24-16.67) | 0.533 | 0.95 (0.09-10.00) | 0.966 |
| 11+ years | 57 (32) | 17 (45) | 0.63 (0.31-1.28) | 0.207 | 0.32 (0.06-1.82) | 0.203 |
| Illness[a]: n(%) | | | | | | |
| Respiratory | 81 (45) | 14 (37) | 1.41 (0.68-2.86) | 0.358 | 0.49 (0.07-3.57) | 0.477 |
| Diarrhoea | 14 (8) | 3 (8) | 0.98 (0.27-3.57) | 0.981 | 0.27 (0.03-2.17) | 0.223 |
| Respiratory and Diarrhoea | 14 (8) | 5 (13) | 0.56 (0.19-1.64) | 0.291 | 0.57 (0.05-7.14) | 0.662 |
| Other | 71 (39) | 16 (42) | 0.89 (0.44-1.82) | 0.761 | Ref | Ref |
| Distance (km)[b]: median (IQR) | 2.6 (1.1-5.9) | 0.9 (0.0-2.3) | **1.37 (1.10-1.72)** | **0.005** | 1.37 (1.06-1.79) | **0.016** |
| Severity (perceived): n(%) | | | | | | |
| Not-Severe | 71 (39) | 20 (54) | Ref | Ref | Ref | Ref |
| Severe | 109 (61) | 17 (46) | 1.82 (0.88-3.70) | 0.104 | 1.54 (0.47-5.00) | 0.480 |
| Time of event[c]: n(%) | | | | | | |
| Day | 83 (49) | 17 (47) | Ref | Ref | Ref | Ref |
| Night | 88 (51) | 19 (53) | 0.95 (0.48-1.92) | 0.901 | 1.11 (0.35-3.57) | 0.865 |
| Transportation: n( %) | | | | | | |
| Informal (foot/ bike/ donkey) | 9 (5) | 1 (3) | **0.33 (0.15-0.71)** | **0.005** | 0.45 (0.01-16.67) | 0.664 |
| Public/ motorcycle taxi | 141 (79) | 23 (61) | **2.44 (1.15-5.00)** | **0.020** | 1.52 (0.06-50.00) | 0.802 |
| Private car/ motorcycle | 29 (16) | 14 (37) | 1.96 (0.24-16.67) | 0.530 | Ref | Ref |
| Highest-education level (HoH): n(%) | | | | | | |
| None | 14 (8) | 8 (21) | **0.32 (0.12-0.82)** | **0.018** | Ref | Ref |
| Below secondary | 69 (38) | 19 (50) | 0.62 (0.31-1.25) | 0.185 | 1.37 (0.31-5.88) | 0.675 |
| Secondary and above | 97 (54) | 11 (29) | **2.86 (1.33-6.25)** | **0.007** | 5.00 (1.05-25.00) | **0.044** |
| Primary source of income: n(%) | | | | | | |
| Agriculture/Animal husbandry | 71 (40) | 15 (43) | 1.11 (0.54-2.27) | 0.782 | NA[e] | 0.991 |
| Vendor (Commerce) | 60 (34) | 10 (29) | 1.54 (0.70-3.33) | 0.283 | NA[e] | 0.992 |
| Tradesperson | 23 (13) | 10 (29) | 0.44 (0.19-1.03) | 0.056 | NA[e] | 0.992 |
| Salaried employment | 23 (13) | 0 (0) | 3.57 (0.46-25.00) | 0.222 | NA[e] | 0.991 |
| Grid cell population density: n(%) | | | | | | |
| Low | 32 (18) | 5 (13) | 1.43 (0.52-4.00) | 0.492 | Ref | Ref |
| Medium | 22 (12) | 11 (29) | 0.34 (0.15-0.79) | 0.011 | 0.54 (0.11-2.56) | 0.440 |
| High | 126 (70) | 22 (58) | 1.69 (0.83-3.45) | 0.149 | 5.56 (1.00-33.33) | **0.050** |
| Events/prior month: median(IQR) | 2 (1-2) | 1 (1-2) | 0.95 (0.63-1.43) | 0.801 | 0.84 (0.43-1.64) | 0.611 |
| S.E.S index[d]: median(IQR) | 3 (2-4) | 3 (2-4) | **2.78 (1.25-6.25)** | **0.012** | (omitted) | (omitted) |

**Footnote for Table S4**

* Total number of providers given (N). Bold text designates p values < 0.05 and 95% CI that do not span 1.00.

[a] Respiratory is defined as 'cough' or 'cold' with fever and no diarrhoea, diarrhoea is with or without blood and no respiratory infection.

[b] Distance calculated as direct distance between household and selected provider.

[c] Day is between 6AM- 6PM and night is between 6PM-6AM.

[d] See methods for variables included in the S.E.S index.

[e] Values not available as model unable to converge.

**Table S5.** Congruence of care-seeking intention and behavior for provider selection

**Table S5.** Congruence of care-seeking intention and behavior for provider selection

| | Care sought from a conventional provider | | | | | Cohen's Kappa *k* (95% CI) | McNemar's OR (95% CI; p value) |
|---|---|---|---|---|---|---|---|
| | Total | StdC+[a] | StdC- | HE+ | HE- | | |
| Illness[b] | | | | | | | |
| Respiratory | 114 (53) | 93 | 9 | 2 | 10 | 0.593 (0.378-0.807) | 0.22 (0.02 – 1.07; 0.065) |
| Diarrhoea | 36 (17) | 27 | 6 | 1 | 2 | 0.276 (-0.092-0.644) | 0.17 (0.00 – 1.37; 0.125) |
| All | 217 | 175 | 27 | 4 | 11 | 0.351 (0.181-0.521) | **0.15 (0.04 – 0.43; <0.001)** |

[a] StdC (standardized case), HE (health event), + (conventional provider), - (non-conventional provider).
[b] Respiratory is defined as 'cough' or 'cold' with fever and no diarrhoea , diarrhoea is with or without blood and no respiratory infection.

**Table S6.** Determinants of non-congruence between intentions and behaviors for provider selection

**Table S6. Determinants of non-congruence between intentions and behaviors for provider selection**

| **Health Events** | All | | OR (CI) | p | aOR (CI) | p |
|---|---|---|---|---|---|---|
| | StdC+/HE+ or StdC-/HE-[a] n=186 | StdC+/HE- or StdC-/HE+ n=31 | | | | |
| Distance: (km)[b] | --- | --- | 0.99 (0.94-1.04) | 0.651 | 1.00 (0.94-1.05) | 0.912 |
| Patient age: n(%) | | | | | | |
| 0-4 years | 116 (64) | 15 (44) | 1.17 (0.82-1.67) | 0.378 | ref | ref |
| 5-10 years | 9 (5) | 1 (3) | 0.78 (0.34-1.78) | 0.552 | 1.63 (0.15-17.22) | 0.685 |
| 11+ years | 57 (31) | 18 (53) | 0.89 (0.61-1.27) | 0.513 | 1.11 (0.26-4.72) | 0.885 |
| Illness[c]: n(%) | | | | | | |
| Respiratory infection | 88 (47) | 8 (24) | 1.19 (0.84-1.67) | 0.326 | 0.31 (0.06-1.78) | 0.191 |
| Diarrhoea | 14 (8) | 3 (9) | 1.15 (0.61-2.17) | 0.677 | 1.19 (0.21-6.68) | 0.843 |
| Respiratory infection and diarrhoea | 15 (8) | 4 (12) | **1.98 (1.10-3.57)** | **0.023** | 0.71 (0.08-6.58) | 0.765 |
| Other | 69 (37) | 19 (56) | **0.57 (0.40-0.82)** | **0.003** | ref | ref |
| Time of event[d]: n(%) | | | | | | |
| Day | 84 (47) | 17 (53) | 0.74 (0.52-1.04) | 0.083 | ref | ref |
| Night | 93 (53) | 15 (47) | 1.35 (0.96-1.91) | 0.083 | 1.00 (0.36-2.83) | 0.997 |
| Transportation: n(%) | | | | | | |
| Informal | 37 (20) | 6 (18) | **1.76 (1.17-2.66)** | **0.007** | 0.36 (0.02-6.23) | 0.479 |
| Public/motorcycle taxi | 139 (75) | 27 (79) | **0.63 (0.42-0.92)** | **0.017** | 0.34 (0.03-4.37) | 0.408 |
| Private car/ motorcycle | 9 (5) | 1 (3) | 0.83 (0.36-1.87) | 0.646 | ref | ref |
| Education: n(%) | | | | | | |
| None | 19 (10) | 3 (9) | 1.61 (0.94-2.75) | 0.085 | ref | ref |
| Below secondary | 70 (38) | 18 (53) | 1.29 (0.91-1.83) | 0.154 | 1.13 (0.23-5.49) | 0.882 |
| Secondary and above | 97 (52) | 13 (38) | **0.63 (0.45-0.89)** | **0.009** | 0.28 (0.05-1.52) | 0.140 |
| Occupation: n(%) | | | | | | |
| Agriculture/Animal husbandry | 79 (45) | 9 (28) | 0.99 (0.69-1.41) | 0.963 | 0 | 0.992 |
| Vendor (Commerce) | 59 (33) | 12 (38) | 0.97 (0.67-1.41) | 0.886 | 0 | 0.993 |
| Tradesperson | 23 (13) | 10 (31) | 1.01 (0.60-1.70) | 0.975 | 0 | 0.993 |
| Employment | 16 (9) | 1 (3) | 0.69 (0.35-1.33) | 0.263 | 0 | 0.992 |
| Population density: n(%) | | | | | | |
| Low | 33 (18) | 6 (18) | 1.15 (0.74-1.79) | 0.526 | ref | ref |
| Medium | 28 (15) | 5 (15) | 1.80 (1.14-2.84) | 0.012 | 1.04 (0.22-5.05) | 0.958 |
| High | 125 (67) | 23 (68) | **0.62 (0.43-0.89)** | **0.009** | 0.26 (0.05-1.35) | 0.110 |
| Events/prior month: median (IQR) | --- | --- | 1.01 (0.83-1.24) | 0.907 | 1.36 (0.73-2.53) | 0.329 |
| S.E.S index[e]: median(IQR) | --- | --- | 0.71 (0.50-1.02) | 0.061 | (omitted) | (omitted) |

**Footnote for Table S6**

[a] StdC (standardized case), HE (health event), + (conventional provider), - (non-conventional provider)
[b] Calculated using straight-line distances.
[c] Respiratory is defined as ‘cough’ or ‘cold’ with fever and no diarrhoea , diarrhoea is with or without blood and no respiratory infection
[d] Day is between 6AM- 6PM and night is between 6PM-6AM.
[e] See methods for variables included in the S.E.S index.

### Household Questionnaire:

*The survey will capture household demographics, socioeconomic status, a recall log detailing health care events with their associated decision making and care seeking behaviors, as well as responses to open ended standardized cases.*

| **Question** | **Format** |
|---|---|
| **Registration** | |
| 1. Household Study Number | Grid (###)-House (###) |
| 2. Date | dd/mm/yyyy |
| 3. Time | _ _ : _ _ (24h) |
| 4. First and last name (head of household) | Free text |
| 5. Primary/ secondary phone number | ####-#### |
| **Phone** | |
| 6. At least one phone has charge now | Yes/No |
| 7. At least one phone has credit now | Yes/No |
| 8. At least one phone is a smart phone | Yes/No |
| 9. Smart phone has data | Yes/No |
| **Location** | |
| 10. Arrondissement | Categorical; drop down |
| 11. Commune | Categorical; drop down |
| 12. Communal section | Categorical; drop down |
| 13. Community | Categorical; drop down |
| 14. GPS location | Latitude/Longitude |
| **Household Resources** | |
| 15. Highest education level of head of household | Categorical; drop down |
| 16. Method of payment for housing (e.g. rent, own) | Categorical; radio |
| 17. Do you own the land that your house is on | Yes/ No |
| 18. Roof material | Categorical; check boxes |
| 19. Wall material | Categorical; check boxes |
| 20. Floor material | Categorical; check boxes |
| 21. Primary water source | Categorical; radio button |
| a) If handpump, where is the nearest pump? | Free Text |
| b) Pump handle sample ID | XX_XX-PH, not taken |
| c) Pump water sample ID | XX_XX-PW, not taken |
| 22. Is this the same primary water source, for drinking? | Yes/no |
| a) If alternative drinking water source, what type? | Categorical; radio |
| 23. Household sanitation method | Categorical; radio |
| 24. Household cooking/fuel source | Categorical; radio |
| 25. Household electricity source | Categorical; radio |
| 26. Household primary/secondary/tertiary transport method | Categorical; radio |
| 27. Animals owned by the household | Categorical; check boxes |
| a) Animals are inside outside the yard | Categorical; radio |
| 28. Sources of household income | Categorical; check boxes |
| 29. Primary source of household income | Categorical; radio button |
| 30. Does household have a bank account? | Yes/ No |
| 31. Immediate family living abroad (mother/child/sibling) | Yes/ No |

**Standardized cases:**

**Case 1:** Respiratory scenario.

*You are awoken at ten at night to the sound of your child coughing…*

| | |
|---|---|
| 32. How would you assess the severity of the cough | Free text |

*The child appears warm when you place your hand on the forehead…*

| | |
|---|---|
| 33. Do you have a thermometer at home? | Yes/ No |
| 34. Do you know what temperatures is considered a fever? | Yes/No |
| a) If yes, what? | XXX (F or C). |
| 35. Do you have paracetamol to treat fever at home? | Yes/ |

*The child does indeed have a fever, is coughing up significant mucus, and is also taking pauses between words when the child speaks. You decide to seek help…*

| | |
|---|---|
| 36. When would you seek help? | Categorical; radio |
| 37. Where would you seek help | Free text |
| 38. What type of provider is this | Categorical; check boxes |
| 39. If permitted, Provider Phone Number | XXXX_XXXX |
| 40. If permitted, Provider Location | Free text |
| 41. What method would you use to contact provider? | Categorical; check boxes |
| 42. Primary/secondary/tertiary reason to choose this provider? | Categorical; check boxes |
| 43. Additional comments about the event | Free text |

**Case 2:** Diarrhea scenario.

*You are awoken at ten at night to the sound of your child asking to go to the bathroom because they have diarrhea…*

| | |
|---|---|
| 44. How would you assess the severity of the diarrhea | Free text |

*The child has 6 loose stools in the last hour…*

| | |
|---|---|
| 45. Do you have ORS (oral rehydration solution at home)? | Yes/ No |
| a) If yes, how much ORS would you give? | Free text |

*The number of stools increases in each hour and it appears watery, like the color of water when you make rice, there is no blood in the stool. You decide to seek help…*

| | |
|---|---|
| 46. When would you seek help? | Categorical; radio |
| 47. Where would you seek help | Free text |
| 48. What type of provider is this | Categorical; check boxes |
| 49. If permitted, Provider Phone Number | XXXX_XXXX; |
| 50. If permitted, Provider Location | Free text |
| 51. What method would you use to contact provider? | Categorical; check boxes |
| 52. Primary/secondary/tertiary reason to choose this provider? | Categorical; check boxes |
| 53. Additional comments about the event | Free text |

**Household Members**

| | |
|---|---|
| 54. Number of household members | ## |
| 55. For each member | |
| a) Age | Categorical; radio |
| b) Sex | Categorical; radio |
| b) Medications taken within the last 10 days | Categorical; check boxes |

If child less than 5 years

| | |
|---|---|
| a) Diarrhea in the last 7 days | Yes/No |
| b) 3+ loose stools in the past 24 hours? | Yes/No |
| c) A bloody stool in the past 24 hours | Yes/No |
| d) Onset less than 7 days ago? | Yes/No |

| | |
|---|---|
| e) Dehydration status | none/some/severe |
| f) Coughing within last 10 days | Yes/No |
| g) Symptoms started within last 10 days | Yes/No |
| h) Fever now or within last 10 days | Yes/No |
| i. 1) If now, measure | ##.# C |
| i) Weight | ##.# kg |
| j) Height | ##.# cm |
| k) MUAC | XXX mm |

**Healthcare Events (recall log)**

For each healthcare event

| | |
|---|---|
| 56. Household member | Categorical; drop down |
| 57. What type of medical problem | Categorical; check box |
| 58. Medical problem description | Optional: free text |
| 59. Severity of problem | Likert scale; radio button |
| 60. How was severity determined? | Free text |
| 61. What was the cause of the problem? | Free text |
| 62. Time problem started | Categorical; radio button # |
| 63. Was it a holiday or weekend? | Yes/ No |
| 64. Was care sought outside the home? | Yes/No |
| a) If no, why? | Categorical; check boxes; |
| b) If treated at home, how? | Categorical; check boxes; |
| c) Outcome | Categorical; check boxes |
| d) Additional comments about the event | Free text |
| If yes… | |
| 65. How many places did you seek care from? | XX |
| 66. If permitted Provider Name | Free text: First, Last |
| 67. If permitted, Provider Phone Number | XXXX_XXXX |
| 68. If permitted Provider Location | Free text; Can refuse |
| 69. What type of provider is this | Categorical; check boxes |
| 70. How was provider contact made? | Categorical; radio |
| 71. Time care was sought | Categorical; radio |
| 72. Was it a holiday or weekend? | Yes/ No |
| 73. Time passed between start of illness and seeking care? | Categorical; radio |
| 74. Why did you choose to seek care when you did? | Free text |
| 75. Primary/secondary/tertiary reason to choose this provider? | Categorical; check boxes |
| 76. What was provider's assessment of severity? | Likert scale; radio button |
| 77. Type of care(s) provided | Categorical; check boxes |
| a) What meds did provider suggest/give | Categorical; check boxes |
| b) What treatment did provider suggest/give | Categorical; check boxes |
| c) What tests did provider suggest | Categorical; check boxes |
| 78. If patient did not obtain meds/treatment- why? | Categorical; check boxes |
| 79. Total cost for consultation/meds/tests/treatment | XXX gourdes |
| 80. Total cost for transportation | XXX gourdes |
| Repeat questions 66 to 80 for each provider visited | |
| 81. What was outcome | Categorical; radio |
| 82. Additional comments about the event | Free text |

### Provider Questionnaire

*The survey will capture general information about conventional and nonconventional healthcare providers, descriptors of their facilities/practices and patients, a recall log detailing cases seen in the last week with the associated decision making process of the provider, as well as responses to open ended standardized cases. Basic contact information about providers will be obtained but no personal information about patients will be obtained.*

| **<u>Question</u>** | **<u>Format</u>** |
|---|---|
| **Registration** | |
| 1. Provider Id | Grid (###)-Facility (#) |
| 2. Date | dd/mm/yyyy |
| 3. Time | _ _ : _ _ (24h) |
| 4. Official name of facility | Free Text |
| 5. Familiar name of facility | Free Text |
| 6. Number of people consented for survey | X |
| For each person | |
| a) Name | First/ Last; free text |
| b) Phone number | (XXXX_XXXX) |
| c) Position at facility | Free text |
| d) Gender | Female/ Male |
| e) Age | Categorical; drop down |
| f) Highest education level completed | Categorical; drop down |
| **Location** | |
| 7. Arrondissement | Drop down menu |
| 8. Commune | Drop down menu |
| 9. Communal Section | Drop down menu |
| 10. Community | Free text |
| 11. GPS location | Latitude/Longitude |
| **Type of Facility/Practice** | |
| 12. Facility has a sign | Yes/No |
| 13. How long has the facility been in operation? | Categorical; Drop down |
| 14. Type of facility (e.g. government, for-profit) | Categorical; check boxes |
| 15. If private, what is monthly income? | Categorical; Drop down |
| 16. What affiliations does the facility have? (e.g. church) | Free text |
| 17. How many employees and what is their structure? | Free text |
| 18. Do employees have appropriate licenses | Categorical; Drop down |
| 19. Are there set hours of operations? | Yes/No; If no skip to #22 |
| 20. What are the days/hours of operation? | Matrix table |
| a) Which days is facility open? | Categorical; Drop down |
| b) What times is the facility open | Categorical; Drop down |
| 21. Do patients contact you at night (6pm+) | Yes/No |
| v. If yes, how many in the past week? | Free text |
| 22. What types of patients do you serve? | Categorical; check boxes |
| 23. Top 3 problems patients < 5 yrs present with | Categorical; Drop down |
| 24. Top 3 problems patients < 5 yrs present with at night | Categorical; Drop down |
| 25. Top 3 problems adult patients present with | Categorical; Drop down |
| 26. Top 3 problems adult patients present with at night | Categorical; Drop down |
| 27. Does the facility have a phone that patients can call? | Yes/No |
| a) If yes, is it charged now? | Yes/No |
| b) Does the phone have minutes now? | Yes/No |

| | |
|---|---|
| c) Is the phone a smart phone? | Yes/No; if no skip to #32 |
| d) Does the phone have data now? | Drop down |
| 28. Does facility have a license to operate | Yes/No |
| 29. What is monthly case load | Free text |
| 30. Do you refer patients to other facilities/providers? | Yes/No |
| a) If yes, give up to 3 referrals | Free text |

**High volume (>200/month) healthcare providers resources/services**

| | |
|---|---|
| 31. What specialties does the facility offer? | Categorical; check boxes |
| 32. Does the facility have a laboratory? | Yes/No |
| 33. Does the facility have a pharmacy? | Yes/No |
| 34. Does the facility provide in-patient services? | Yes/No |
| 35. Does the facility have an ambulance service? | Yes/No |
| 36. Additional comments/ questions | Free text |

**Low volume (<200/month) healthcare providers resources/services** XXXX

| | |
|---|---|
| 37. Have you taken university courses related to business? | Yes/No |
| a) If yes, explain | Free text |
| 38. Have you attended trainings related to your business? | Yes/No |
| a) If yes, explain | Free text |
| 39. Is there a designated structure for the facility | Yes/No |
| 40. Facility payment method (e.g. rent, own) | Categorical; radio |
| 41. Roof material | Categorical; check boxes |
| 42. Wall material | Categorical; check boxes |
| 43. Of the last 10 patients how many were strangers? | ## |
| 44. Most frequent methods patients contact you? | Categorical; check boxes |
| 45. What type of care do you provide? | Categorical; check boxes |
| 46. What equipment/resources do you have? | Categorical; check boxes |
| 47. Do you provide ORS? | Yes/No |
| a) If yes, what type? | Categorical; radio |
| b) What is the cost? | ### gourdes per unit type |
| 48. Are you contacted by patients with respiratory illness? | Yes/No |

**Standardized cases**

**Case 1:** Respiratory scenario.

*If yes, you are contacted at ten at night by cell phone from a mother who says her 2-year old child has cough…*

| | |
|---|---|
| 49. How would you assess the severity of the cough? | Free text |

*The mother says the child appears warm when she places her hand on the child's forehead…*

| | |
|---|---|
| 50. The mother has a thermometer, what temp is a fever? | XXX (F or C). |
| 51. If the child needs paracetamol, how much would you prescribe? | Free text |

*The child does indeed have a fever, is coughing up significant mucus, and is also taking pauses between words when the child speaks.*

| | |
|---|---|
| 52. What do you recommend the mother do? | Categorical; radio |
| 53. Why did you make this recommendation | Free text |
| a) If treat at home, how? | Free text |

| | |
|---|---|
| b) If seek care, when? | Categorical; radio |
| c) Where should the patient go? | Free text |
| d) If permitted, referral provider phone number | XXXX_XXXX |
| e) If permitted, referral provider location | Free text |
| f) Why did you make this recommendation? | Free text |
| g) What type of provider is this? | Categorical; Drop down |
| h) What type of care should this patient get? | Categorical; Drop down |
| i) What meds should patient get? | Categorical; check boxes |
| j) What treatment should patient get? | Categorical; check boxes |
| k) What tests should patient get? | Categorical; check boxes |
| l) How much would this care cost? | ### gourdes |
| m) Additional comments about the event | Free text |
| 54. Are you contacted by patients with diarrhoeal illness? | Yes/No |

**Case 2:** Diarrhea scenario.

*You are contacted at ten at night by cell phone from a mother who says her two--year old child is asking to go to the bathroom because of diarrhea…*

55. How would you assess the severity of the diarrhea? — Free text

*The child has 6 loose stools in the last hour…*

56. The family has ORS at home, how much would you recommend? — Free text

*The number of stools increases in each hour and it appears watery, like the color of water when you make rice, there is no blood in the stool. You recommend…*

| | |
|---|---|
| 57. What do you recommend the mother do? | Categorical; radio |
| 58. Why did you make this recommendation | Free text |
| a) If treat at home, how? | Free text |
| b) If seek care, when? | Categorical; radio |
| c) Where should the patient go? | Free text |
| d) If permitted, referral provider phone number | XXXX_XXXX |
| e) If permitted, referral provider location | Free text |
| f) Why did you make this recommendation? | Free text |
| g) What type of provider is this? | Categorical; Drop down |
| h) What type of care should this patient get? | Categorical; Drop down |
| i) What meds should patient get? | Categorical; check boxes |
| j) What treatment should patient get? | Categorical; check boxes |
| k) What tests should patient get? | Categorical; check boxes |
| l) How much would this care cost? | ### gourdes |
| m) Additional comments about the event | Free text |

**Provider Healthcare Events (recall log)**

59. In the past week how many times has someone sought healthcare services from you? — ##

For each health event

| | |
|---|---|
| 60. Patient sex | Female/ Male |
| 61. Patient age | Categorical; dropdown |

| | |
|---|---|
| 62. Health event type | Categorical; dropdown |
| 63. Health event description | Free text |
| 64. Severity of problem | Likert scale; radio button |
| 65. How was severity determined? | Free text |
| 66. What do you think caused the problem? | Free text |
| 67. What time did patient contact you? | Categorical: dropdown |
| 68. Was it weekend/weekday/holiday | Categorical: radio |
| 69. How did patient contact you? | Categorical; check boxes |
| 70. Was treatment provided? | Yes/No |
| 71. What treatment was provided? | Categorical; check boxes |
| 72. What medication was provided? | Categorical; check boxes |
| 73. What tests were provided? | Categorical; check boxes |
| 74. How much time passed between illness onset | |
| 75. and contacting you? | Categorical; radio |
| 76. What was cost of treatment? | XXXX gourdes |
| 77. Time treatment was provided? | Categorical; radio button |
| 78. Was patient referred? | Yes/No; |
| a) If yes, where was patient referred? | Free Text |
| b) If permitted, referral provider phone number | XXXX_XXXX |
| c) If permitted, referral provider location | Free text |
| d) What type of provider is this? | Categorical; dropdown |
| 79. Outcome for patient | Categorical; check boxes |
| 80. Additional comments | Free text |
